# Causal relationship between Environmental Enteric Dysfunction (EED), WaSH practices and growth failure in children from Rukwa-Tanzania

**DOI:** 10.1101/2022.02.04.22270429

**Authors:** Grantina Modern, Emmanuel Mpolya, Elingarami Sauli

## Abstract

This was a hospital based prospective cohort study aimed at assessing causal relationship between environmental enteric dysfunction (EED), WaSH practices and growth failure in children aged between 1-2 years from Rukwa, Tanzania, using univariate and multivariate logistic regression models. We found very significant causal relationship between growth failure and combined water availability (p=0.0055), toilet sharing by parents/caregivers (OR= 2.632, CI: 1.156 - 6.233, p=0.023 from univariate analysis, and OR=4.067, CI: 1.484 - 12.206, p=0.008 from multivariate analysis), and handwashing by parents/caregivers before food (OR=3.363, CI: 0.928 -11.454, p=0.05 from univariate analysis and OR=15.038, CI:2.422 - 115.8, p=0.005 from multivariate analysis). This significant causal relationship may be linked to increased diarrheal incidence and intestinal worm infections among the studied children, which may then lead to inadequate absorption of nutrients and subsequent growth failure. The findings herein should therefore improve policy and programs designed to deliver targeted intervention strategies in Tanzania, to also identify and include other entry points for managing child growth failure and improving WaSH practices.

## 1. INTRODUCTION

Undernutrition affects 25% of children in the developing world and has been associated with half of all deaths worldwide. The known forms of chronic undernutrition include; wasting (low weight for height), stunting (low height for age), underweight (low weight forage) and deficiencies in vitamins and minerals (1). Stunting is the most common and prevalent form of chronic undernutrition (2) that affects approximately 165 million children globally, equivalent to 23% of all children below the age of five (3). Rukwa had highest prevalence of stunting in Tanzania (56%) among children aged between 1-2 years in 2015 (4), despite it being one of the regions in Tanzania with secured food production.

The long-term effects of stunting in early childhood have been associated with cognitive and physical growth deficits across generations (5). Models depicting use of all known interventions to tackle under nutrition, including vitamin A and zinc supplementation, complementary feeding, breastfeeding promotion and micronutrient supplementation in pregnancy, have shown that their use in 99% of children would only decrease stunting by 33% worldwide, clearly indicating that there is a large knowledge gap in our understanding of stunting and its associated causes that cannot only be explained by food insecurity (6). The main objective of this study was therefore to assess causal relationship between Environmental Enteric Dysfunction (EED), WaSH practices and growth failure among children from Rukwa, Tanzania.

## 2. MATERIALS AND METHODS

### 2.1. Study design, sample size and sampling design

This was a hospital-based prospective cohort study to assess causal relationship between EED, water, sanitation and hygiene (WaSH) practices, and growth failure in children below 2 years of age from Rukwa, Tanzania.

Two health care facilities; Mazwi RCH Clinic and Kizwite Health Care Centre were conveniently selected as data collection centers to equally represent caretakers/parents from households (HHs) from poor versus improved environmental conditions respectively. Children were then randomly selected from the two healthcare facilities to make a sample of 149 children below two years of age. This sample size was calculated using the Kothari 2004 formula (7); which is used for cross-sectional survey. In this formula, the stunting prevalence of 56% for Rukwa (4) was used to calculate a sample size at 5% a precision for rejecting a true null hypothesis, as well as z-score of 1.96 for the 95% confidence interval. This gave us a formula with enough power to show true differences in proportions of growth failure based on covariates that were statistically significantly causing such a difference. Since the population of Sumbawanga (Rukwa) is more than 10,000, no correction for finite population was deemed necessary, and the response rate was 100%. Children below two years of age without medical complications, and whose mothers were willing to participate were included in this study. Those above the stated age were excluded along with children below two years of age who were sick.

### 2.2. Socio-demographic, anthropometric and WaSH data collection

A structured questionnaire with clearly defined and harmonized questions was used in this study to obtain information on socio-demographic characteristics of the study population, anthropometric data (for nutritional status determination), water availability, hygiene and sanitation (WaSH) practices, as well as occurrence of infections. The questionnaire was deployed twice during a six months’ follow up period in 2021. The data was collected at the beginning of follow up (to give baseline information) and at the end of the six months follow up. The questionnaire was administered at the named data collection centers through interviews to parents/guardians of selected children. Height/length (in cm) was measured using the UNICEF height/length board to a precision of 0.1 cm, while weight (in Kg) was measured using a calibrated electronic scale to a precision of 0.1 kg. Determination of nutritional status (stunting, underweight, and wasting) of each child was done using WHO growth standards (Z-scores) and Tanzania Nutrition Assessment, Counseling and Support (NACS) job Aids of 2016.

### 2.3. Urine and stool samples collection

Urine and stool samples were collected from each child using universal bottles. The samples were collected only once for each child. Prior to urine collection, a 20 mLs disaccharide solution (a mixture of Lactulose and Mannitol sugars) was given to the selected children, followed by intake of 300 mLs of drinking water. Urine samples were then collected after 30 minutes to two hours, for lactulose-mannitol ratio test. Collected urine samples were then put into aliquots in 2 mLs cryovials, mixed with three drops of Thimerosal as preservative and stored in a -20°C freezer, awaiting transportation to NM-AIST laboratory for analysis. Stool samples were randomly collected for fecal myeloperoxidase (MPO) marker investigation. These samples were mixed thoroughly and put into aliquots in 2 mLs cryovials, then by stored in a -20°C freezer also awaiting transportation to NM-AIST laboratory for analysis.

### 2.4. Laboratory sample analysis

Determination of EED through the Lactulose: Mannitol (L:M) and MPO test was conducted at NM-AIST laboratories. EED was characterized in this study by increased gut permeability and inflammation, as they are associated with linear growth faltering in children living in low-income countries.

In the L: M test, both lactulose and mannitol were excreted intact by children in the urine after oral ingestion, following minimal metabolism. Urinary mannitol therefore gave us an index of absorptive capacity, while lactulose in the urine indicated impaired barrier function. Higher urinary L: M ratios thus reflected greater abnormalities of one or both functions. Data also suggest that fecal myeloperoxidase (MPO) is a useful test for gut inflammation and that it also predicts linear growth deficits. Prior to the assays, the sample aliquots and sample analysis kits were equilibrated to room temperature as per Manufacturer’s instructions. All assay procedures for L:M ratio and MPO were conducted according to the manufacturer’s protocols. Optical densities were then read at 565 nm using an UV spectrometer and results were recorded in the form of an excel sheet.

### 2.5. Data analysis

Data entry and processing was done manually into an Excel sheet, which was later cleaned, arranged and coded for analyses. The cleaned and coded data was then imported and attached into the R script of R software version 3.4.4 and ran for descriptive statistics. Univariate and multivariate analyses (linear regression models) were used to assess the risk (causal relationship) posed by individual EED markers (MPO and L: M ratios) and WaSH practices in causing growth failure/stunting. From the model summaries, odds ratios, their 95% confidence intervals and p values were extracted to form summarized tables of results for discussion. Significance was considered at p < 0.05 for all statistical analyses.

### 2.6. Ethical Approval

This study was approved by the Northern Tanzania Health Research Ethics committee (KNCHREC), with approval number KNCHREC0011.

## 3. RESULTS AND DISCUSSION

The general objective of this study was to assess the causal relationship between environmental enteric dysfunction (EED), WaSH practices and growth failure in children aged between 1-2 years from Rukwa (Sumbawanga District)-Tanzania. Rukwa was chosen for this study due its highest prevalence of stunting (56%) during the study period (4). Also, Rukwa is known in Tanzania for having secured food production, and yet it had the highest prevalence of stunting. This raised the hypothesis for this study that, perhaps growth failure and stunting among children in Rukwa may also be associated with other factors apart from commonly known food insecurity.

We used a structured questionnaire with clearly defined and harmonized questions to obtain information on socio-demographic characteristics of the study population, nutritional status, water availability, hygiene and sanitation (WaSH) practices, as well as occurrence of infections/morbidity. The structured questionnaire was deployed twice during the study, at the beginning of follow up (to give baseline information) and at the end of the follow-up. The questionnaire was administered at data collection centers through interviews to parents/guardians of the selected children. Two data collection Centers were set, one representing the urban sub population that attended the Mazwi health care Centre, while the other represented rural sub population that attended Kizwite clinic.

## 3.1. Results

Table 1 below represents descriptive statics for socio demographic characteristics, nutritional status, EED, and WaSH practices of the study population.

**Table 1:**
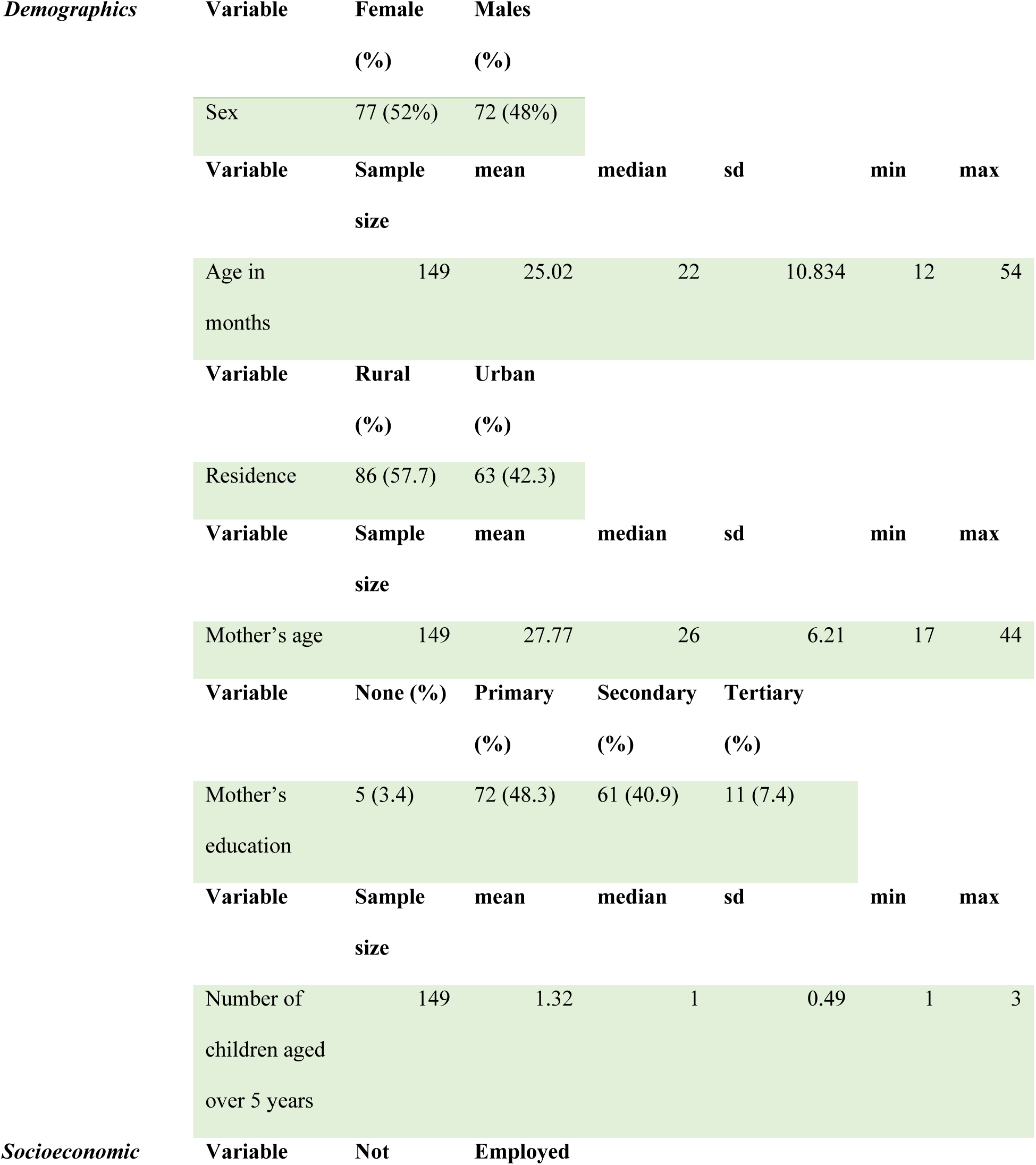
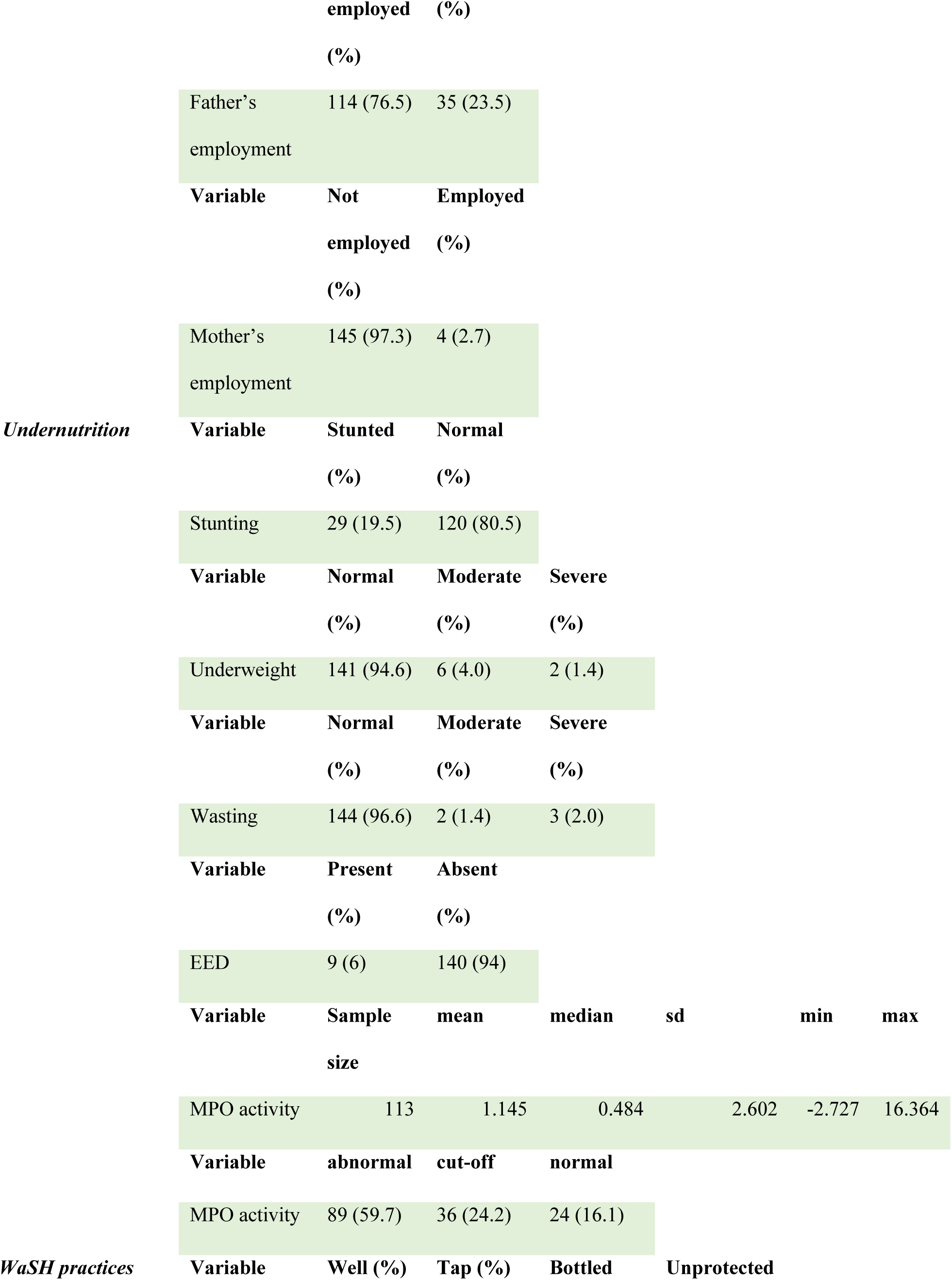
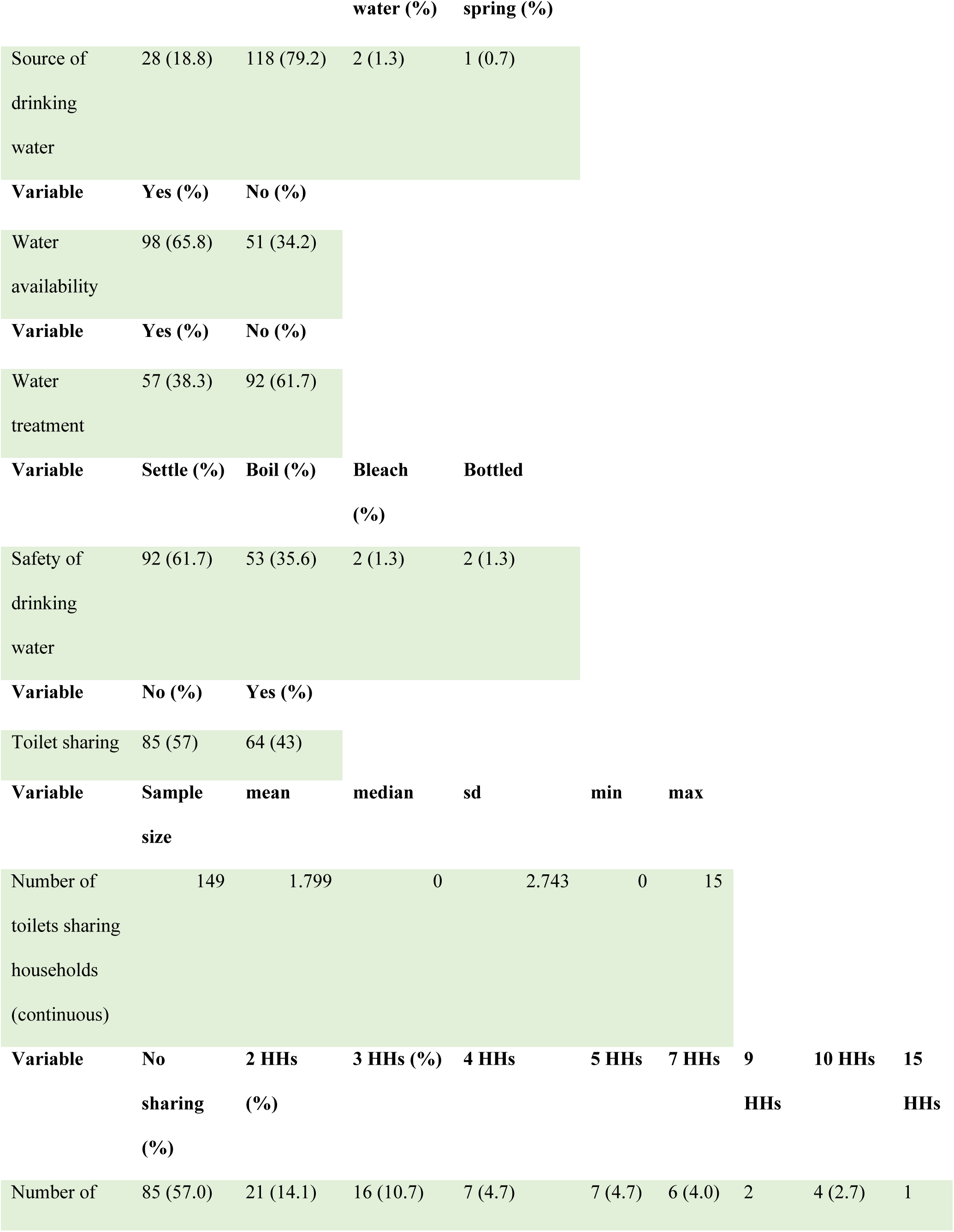
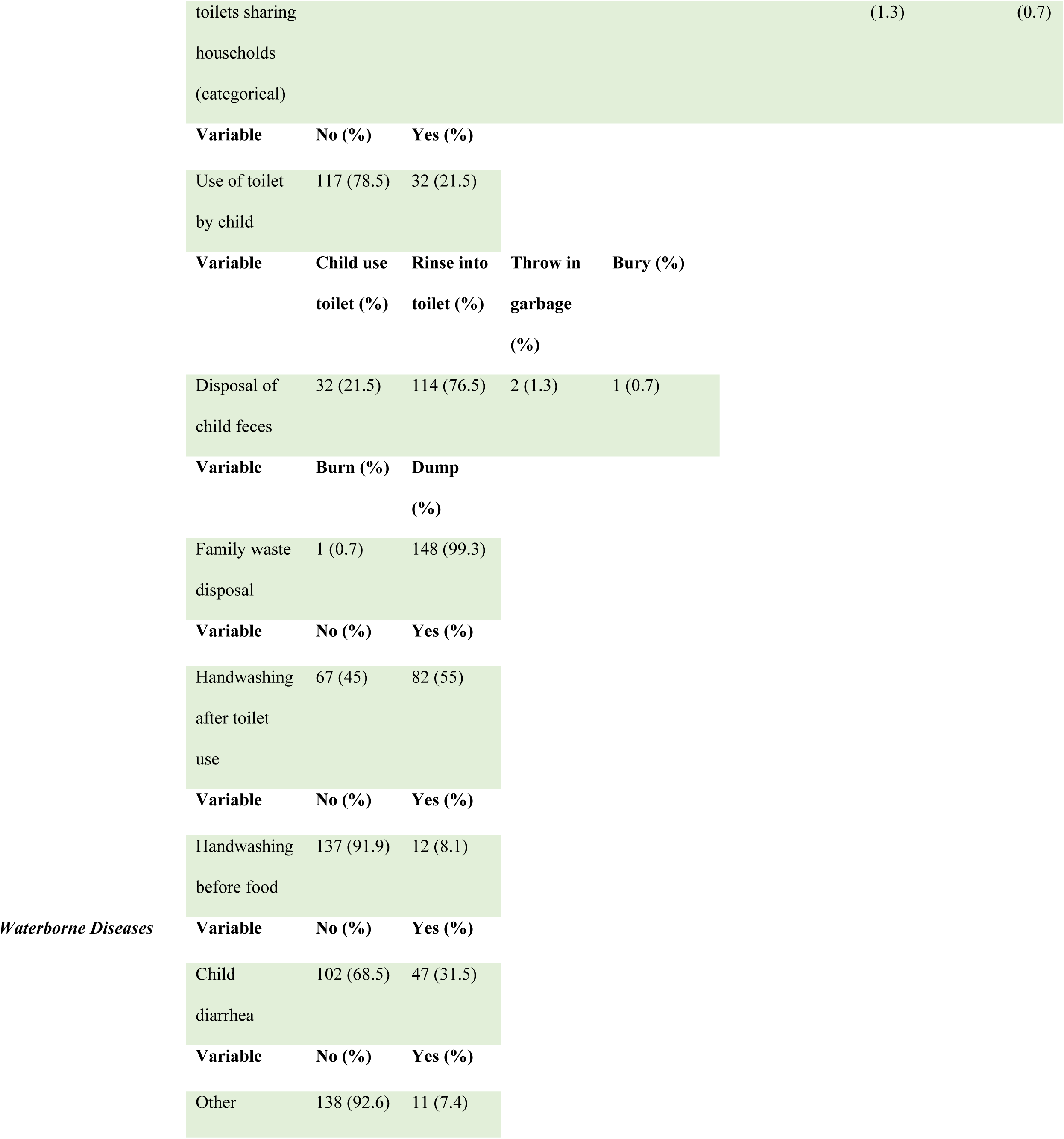
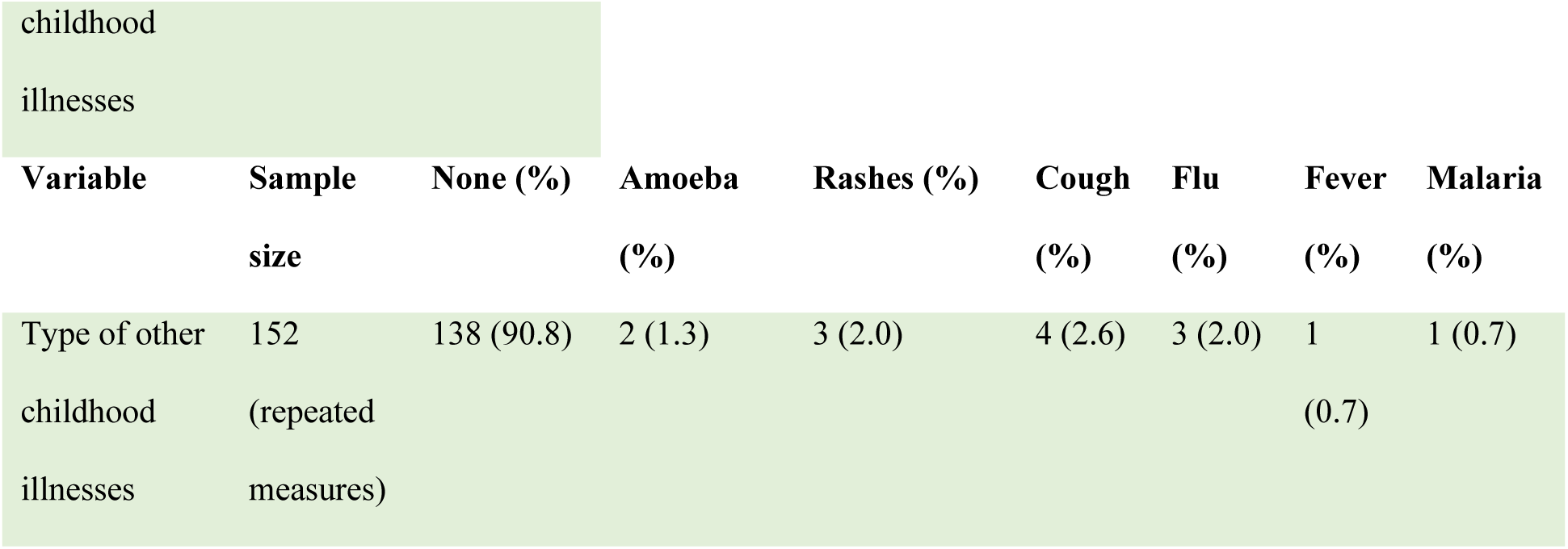
Descriptive statistics for socio demographic characteristics, nutritional status, EED, and WaSH practices of study population

Overall, there were 149 children recruited for this study, of whom 77 (52%) were females and 72 (48%) were males. The children age ranged between 1-2 years, with 86 (57.7%) of them from the rural attending the Kizwite health care Centre, while 63 (42.3%) from urban attended the Mazwi RCH clinic. The children’s mean mother’s age was 27 as seen from table 1, which shows that the mothers’ age was generally young. The children’s parents were hugely unemployed, with 114 (76.5%) unemployed fathers and 145 (97.3%) unemployed mothers, which may have a negative impact in decision making on matters related to their children’s health care. Moreover, there were generally lower levels of undernutrition as seen from table 1, with only 29 (19.5%) stunted cases, 6 (4%) moderate underweight cases, 2 (1.4%) severe underweight cases, 2 (1.4%) moderate wasting, and 3 (2%) severe wasting cases.

As for EED, only 9 (6%) of children were found to have EED, while 140 (94%) of remaining children were found to have no EED. However, there was increased MPO activity in 89 (59.7%) of children, while only 24 (16.1%) of children had normal MPO activity (cut-off: 36 (24.2%).

As for WaSH practices, 118 (79.2%) of children used tap water while 28 (18.8%) consumed water from the wells, and the rest used water from other sources. Overall, only 57 (38.3%) of children consumed treated/boiled water while the rest 92 (61.7%) consumed un treated/un boiled water, putting them at major risk of getting waterborne diseases. As for toilet use, majority of caretakers/parents 117 (78.5%) did not use toilet while only 32 (21.5%) used toilet, this also putting majority of them at high risk of contaminating their children, which leads to frequent diarrhea and other waterborne infections. Moreover, 137 (91.9%) of parents/caretakers did not wash their hands before food and only 12 (8.1%) of them did wash their hands before food. As for toilet sharing, 64 (43%) of caretakers/ parents shared their toilet while 85 (57%) did not share their toilet. There were only 47 (31.5%) reported cases of diarrhea among the studied children, with remaining 102 (68.5%) having no reported diarrhea.

The risk posed by individual EED markers and other studied factors in causing growth failure/stunting was in this study determined by univariate and multivariate logistic regression models as depicted in table 2 below.

**Table 2:**
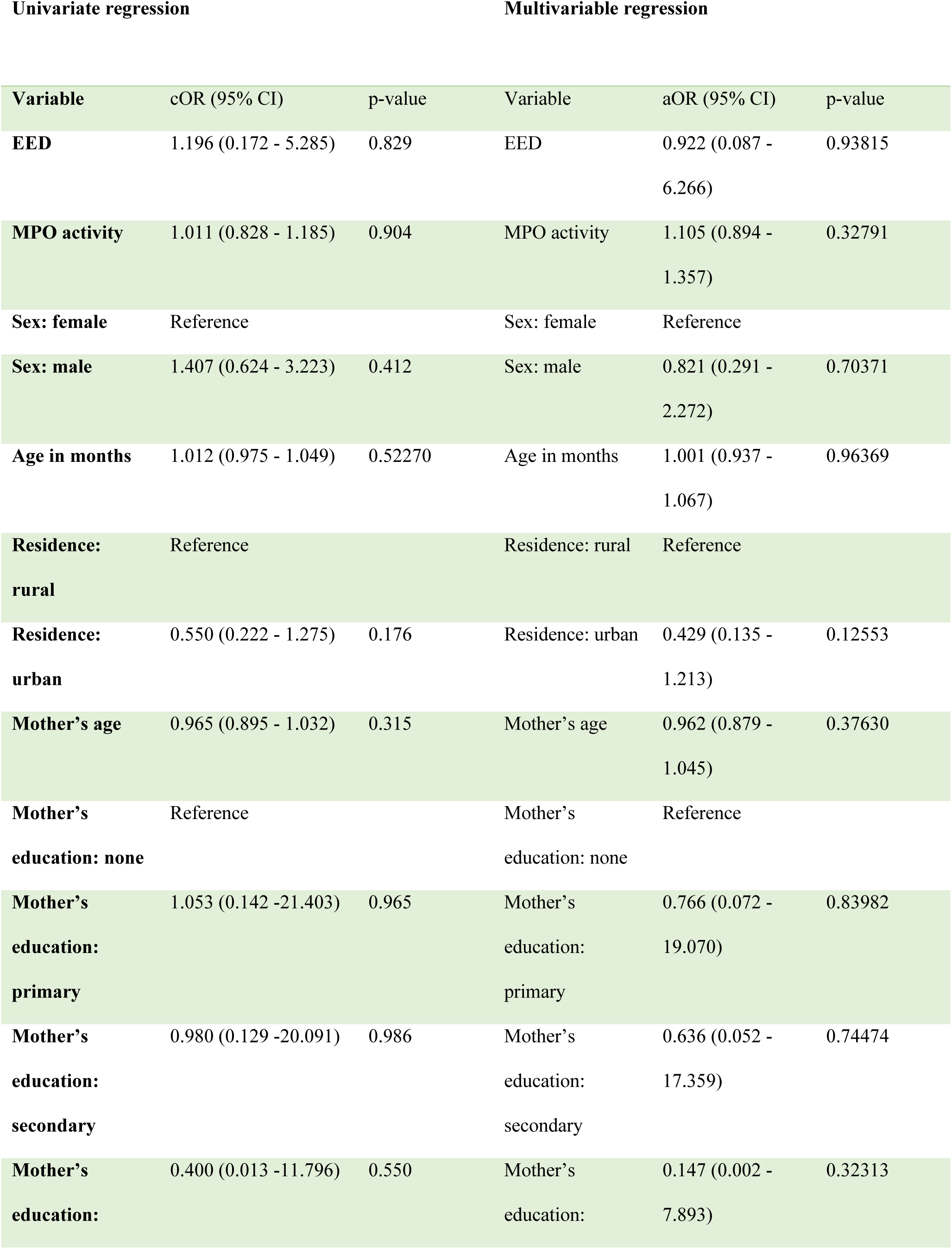
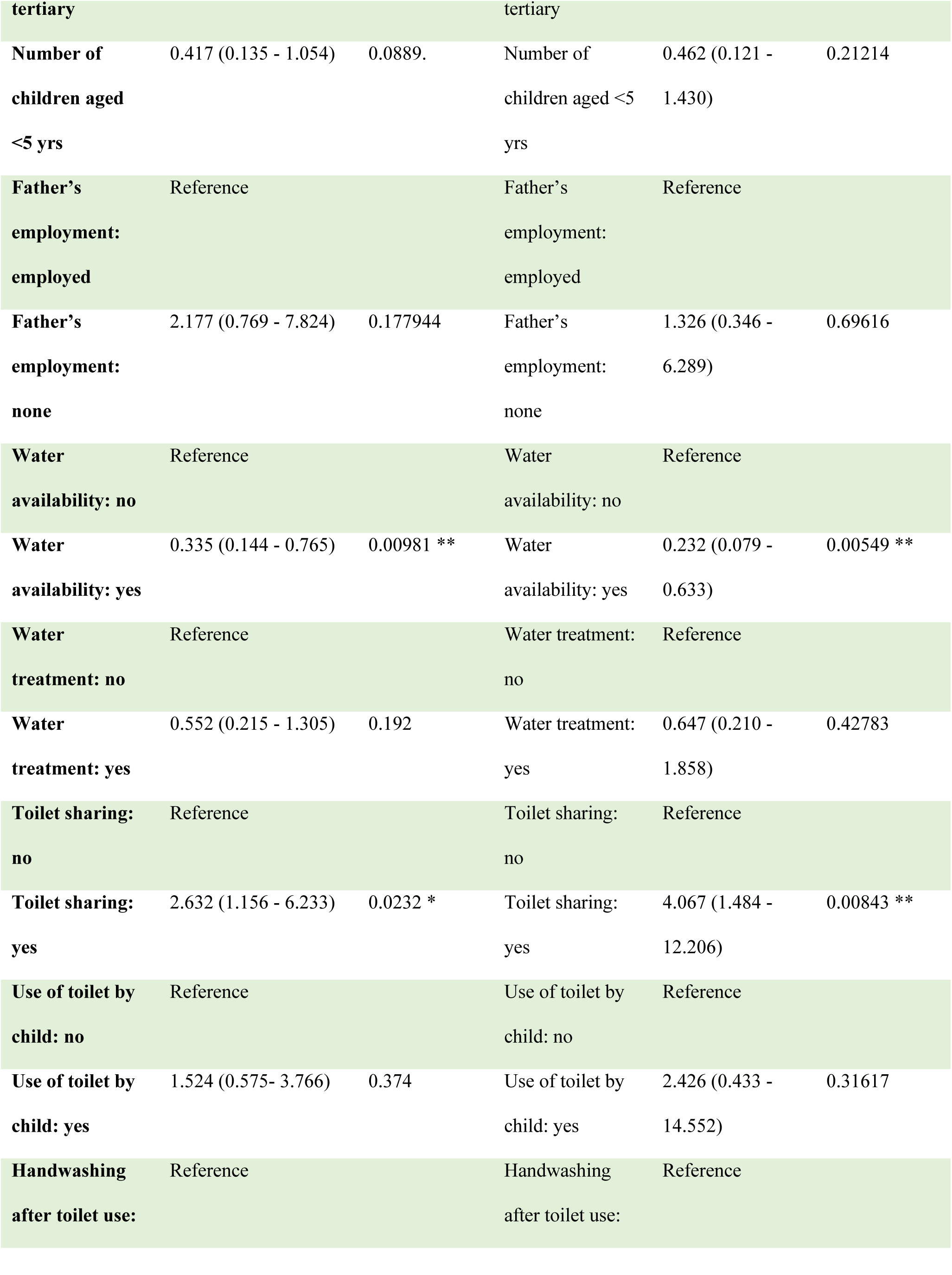
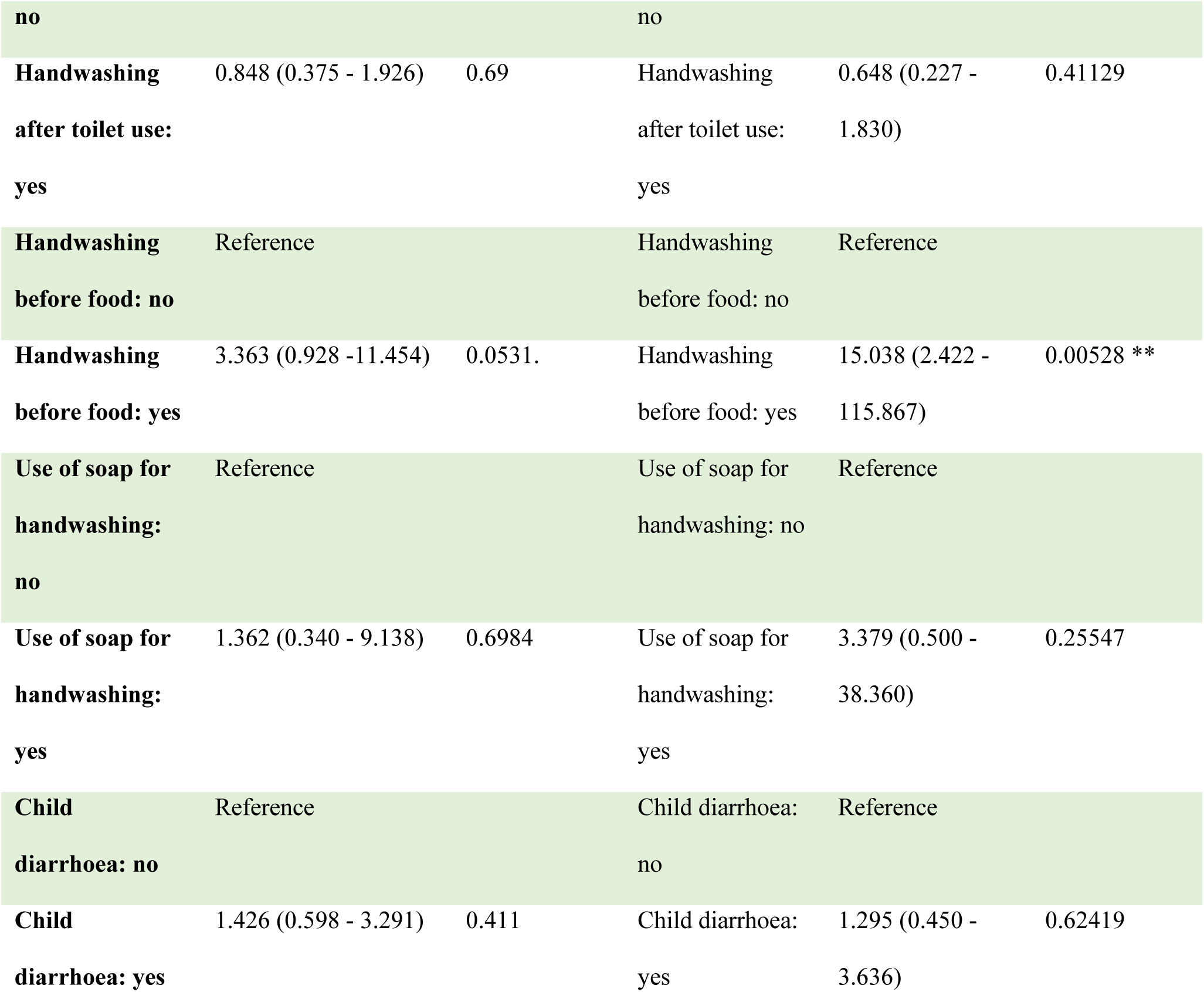
Univariate and multivariate logistic regression models’ output for predictors of stunting/linear growth deficits

As seen from table 2, there was no causal association between growth failure/stunting and the following factors/variables; EED, MPO activity, age of child (in months), residence, mother’s age, mother’s education, number of children, father’s employment, water treatment, use of toilet by child, hand washing after toilet, use of soap for hand washing, and child’s diarrhea. However, there was very significant causal relationship between stunting/growth failure and water availability, with p=0.0098 from univariate analysis and p=0.0055 from multivariate analysis. Moreover, there was significantly increased risk between growth failure/stunting and toilet sharing among parents/caretakers (OR=2.632, CI: 1.156 - 6.233, p=0.023 from univariate analysis and (OR=4.067, CI: 1.484 - 12.206, p=0.008 from multivariate analysis) as seen from table 2.

Handwashing before food by parents/caretakers was also found to have very significant high risk (OR=3.363, CI: 0.928 -11.454, p=0.05 from univariate analysis and OR=15.038, CI:2.422 - 115.8, p=0.005 from multivariate analysis) of causing growth failure/stunting among the studied children in this study, as seen in table 2.

## 3.2. Discussion

A multivariate logistic regression model was in this study applied to assess combined causal association between access to WaSH practices and child growth outcomes in Rukwa, Tanzania, and odds ratios (ORs) were estimated as indicated in table 2. We found very significant causal relationship between stunting/growth failure and water availability (p=0.0098 from univariate analysis and p=0.0055 from multivariate analysis). This significant causal relationship may be due to water being the main link to growth failure through its use in hand washing and cooking, lack of which may lead to frequent contamination and diarrhea and subsequent growth failure among affected children. Another link between water and growth failure/stunting may be through consumption of un treated/un boiled water as seen by majority of children in this study (92 (61.7%)).

Moreover, there was significantly increased risk between growth failure/stunting and toilet sharing among parents/caretakers as seen from table 2 (OR= 2.632, CI: 1.156 - 6.233, p=0.023 from univariate analysis and (OR=4.067, CI: 1.484 - 12.206, p=0.008 from multivariate analysis). This may be due to use of contaminated water during toilet sharing by caretakers or use of shared contaminated toilet by caretakers, which may also lead to frequent contamination of their children, which may then lead to frequent diarrhea and subsequent growth failure. A study by Bekele. T. et al (8) found that, combined access to improved water, sanitation and handwashing was associated with reduced child linear growth failure in Ethiopia.

Handwashing before food by parents/caretakers was also found to have very significant high risk of causing growth failure/stunting among the studied children in this study (OR=3.363, CI: 0.928 -11.454, p=0.05 from univariate analysis and OR=15.038, CI:2.422 - 115.8, p=0.005 from multivariate analysis) as seen in table 2. This may again be directly linked to frequent direct contamination of children (by their caretakers) during feeding, which may then cause frequent waterborne infections/diarrhea among the children in the study population.

Safe drinking water, effective sanitation and adequate hygiene (WaSH) services are key drivers of human health and nutrition (9). Freeman and colleagues estimated in 2014 that, only 19% of the world’s population washed their hands after contact with excreta and only 14% of people in Sub-Saharan Africa wash their hands with soap after using toilet or before eating (10). Inadequate food intake, frequent infections, micronutrient deficiencies, and poor WaSH practices have all been found to contribute to child growth failure. The significant association between child growth failure, poor access to clean water, handwashing by parents/caretakers before food, and poor toilet facilities in this study may be linked to increased diarrhea incidence (11) and intestinal worm infections, which lead to inadequate/poor absorption of nutrients and subsequent growth failure of children (12). Strategies have been put in place in Tanzania to reduce child growth failure through nutritional interventions and diarrheal disease control (13)(14). However, there has been limited knowledge on the extent to which inadequate/poor WaSH practices have been contributing to child growth failure in Tanzania, which was the main focus of this study.

To quantify the causal association between environmental enteric dysfunction (EED) and linear growth failure in children from a region with highest prevalence of stunting in Tanzania (Rukwa), we also assessed myeloperoxidase (MPO) and L:M ratios. Data suggest that fecal myeloperoxidase (MPO) is a useful test for gut inflammation and it also predicts linear growth deficits (15). Myeloperoxidase (MPO) is an enzyme found in neutrophils (16) and, at much lower concentration in monocytes and macrophages (17)(18). This enzyme catalyzes the reaction of hydrogen peroxide and halide ions to produce cytotoxic acids, such as hypochlorous acid (19). The level of MPO in a suspension of neutrophils was found in another study to be directly proportional to the number of neutrophils present over a wide range of neutrophil concentrations. Stool MPO levels were also found to be significantly higher in patients with active inflammatory bowel disease (IBD) compared to patients with inactive disease and normal controls. The MPO activity was also present in low amounts in the stool of patients with inactive IBD and normal controls, and was markedly increased in patients with active disease (20). Although we found increased MPO activity in the studied children in this current study (table 1), the increased MPO activity was not significantly related to growth failure/stunting (table 2), independent of whether the children were from rural or urban environments, and this was also the same for EED, which was found in a limited number of children from both urban and rural environments as seen in table 1. However, despite the findings from this current study, other bigger epidemiological studies have found increased levels of inflammation (as measured by individual biomarkers like MPO and EED) associated with growth failure outcomes in most diverse epidemiologic settings, suggesting that the patterns of intestinal injury, or at least those assessed with these markers are consistent in diverse epidemiologic contexts in children living in poverty like Tanzania. Study findings by Kosek. M et al; (2013) (15) also suggested that intestinal inflammation is an important component of environmental enteropathy. Linear growth failure/stunting is a known persistent problem in children living in poverty in the developing world, including Tanzania, hence we propose conduction of bigger epidemiological studies that will measure intestinal inflammation level as a composite disease activity score rather than individual biomarkers, to give proper account of growth failure pattern and monitoring of other possible inflammatory conditions like IBD in children.

## 4. CONCLUSIONS

We in this study found significant causal association between growth failure and combined water availability, hand washing before food and toilet sharing by parents/caretakers in Rukwa. Future research with robust methods is therefore needed to examine whether these combined WaSH practices have synergistic effect on child growth outcomes in Tanzania. The proposed study should also measure intestinal inflammation level as a composite disease activity score rather than individual biomarkers, to give proper account of growth failure pattern and monitoring of other possible inflammatory conditions like IBD in children. The findings herein should also improve policy and programs designed to deliver targeted intervention strategies, to also identify and include other entry points for child growth failure and WaSH practices.

## Data Availability

All relevant data are within the manuscript and its Supporting Information files.

## Acknowledgements

We acknowledge the Sumbawanga (Rukwa) District Council, NM-AIST Management and laboratory, for ensuring smooth implementation of this research work despite all the incurred challenges. This research work was fully supported by a Grant from the Nestle Foundation for the study of problems of nutrition in the World, Lausanne, Switzerland.

## Authors’ contributions

**Grantina Modern:** Research conception, proposal development, field and lab work, and manuscript writing

**Emmanuel Mpolya:** Statistical analysis

**Elingarami Sauli:** Research conception, proposal development and implementation, field and lab work supervision, manuscript writing and revision.

